# Detection of patient metadata in published articles for genomic epidemiology using machine learning and large language models

**DOI:** 10.1101/2025.04.25.25326298

**Authors:** Ari Z. Klein, Davy Weissenbacher, Karen O’Connor, Amir Elyaderani, Ivan Flores Amaro, Takeshi Onishi, Su Golder, Kaelen Spiegel, Matthew Scotch, Graciela Gonzalez-Hernandez

## Abstract

**Objective:** Patient metadata exist in published articles, but are often disconnected from genome sequences in databases, limiting their utility for genomic epidemiology. The objective of this study was to develop and evaluate natural language processing methods to facilitate the large-scale detection of patient metadata associated with reports of genome sequencing in published articles, drawing on the case of SARS-CoV-2.

**Methods:** We applied filters to select a sample of 245 PubMed articles (50,918 sentences) in LitCovid for manual annotation of sentences that reported generating SARS-CoV-2 sequences. We trained, deployed, and validated a BERT-based classifier, and selected a sample of 150 predicted articles (22,147 sentences) for manual annotation of sentences that reported patient metadata associated with the sequences. In addition to training BERT-based classifiers, we experimented with a generative AI approach, prompting the Llama-3-70B LLM using zero-shot, role-based, few-shot, chain-of-thought, and reasoning-eliciting prompting.

**Results:** BERT-based models that were pre-trained on corpora in biomedical or, more specifically, COVID-19 domains outperformed those that were pre-trained on corpora in general domains for detecting reports of patient metadata associated with SARS-CoV-2 sequences, achieving the best performance with a classifier based on a BiomedBERT-Large-Abstract model (F_1_-score = 0.776). While the best performance of our generative AI approach was achieved using role-based, few-shot, and chain-of-thought prompting (F_1_-score = 0.558), it was nonetheless outperformed by all of our machine learning-based classifiers.

**Conclusion:** Our methods were applied to more than 350,000 published articles and can be used to advance the utility and efficiency of genomic epidemiology for public health responses to virus outbreaks.

## 1. Introduction

Genomic epidemiology—that is, the use of genome sequences to understand the transmission patterns of viruses—depends on metadata such as the patients’ age, sex, geographic location, travel history, symptoms, and clinical outcomes [1]. While millions of SARS-CoV-2 genome sequences from patients worldwide have been digitally deposited in public databases such as GISAID [2] and GenBank [3], the patient metadata associated with the sequences are often not available in the databases [4, 5]. The COVID-19 pandemic brought to attention this two-fold problem: (1) such metadata, or “dark data,” exist in published articles but are disconnected from digital resources, and (2) manually extracting data from the unstructured text of thousands of articles would be inefficient for timely public health responses to virus outbreaks [6]. Our prior work has demonstrated that host geographic locations associated with genome sequences are available in PubMed articles and can be automatically extracted from unstructured text using natural language processing (NLP) methods [7, 8]. Building on this work, the objective of the present study was to develop and evaluate NLP methods to more broadly facilitate the large-scale detection of various types of patient metadata associated with reports of genome sequencing in published articles, drawing on the case of SARS-CoV-2.

## 2. Methods

### 2.1. Data collection

We downloaded all 210,059 publications that were available at the initial time of this study in LitCovid—a collection of PubMed articles that are automatically identified as relevant to COVID-19 [9]. Our first objective was to develop a high-recall approach for detecting the subset of articles that reported generating SARS-CoV-2 sequences. We developed and applied a series of filters to select a sample of articles to manually annotate for training and evaluating machine learning methods. First, we searched the publications for a “References” section to remove those that included only an abstract without a full-text article, resulting in 88,044 (41.9%) of the articles in LitCovid. Next, we excluded those that mentioned “RNA-Seq” because our focus is on genomic epidemiology rather than gene expression, resulting in 86,604 (41.2%) of the articles in LitCovid. Then, we reviewed a random sample of 100 of them to develop and search for a list of keywords generally related to sequencing (e.g., *sequences, extracted, synthesized*), matching 26,647 (12.7%) of the articles in LitCovid. Finally, we reviewed another random sample of 100 articles to determine whether they reported generating SARS-CoV-2 sequences, and improved precision by searching for additional keywords related to specific sequencing technology (e.g., *oxford nanopore, minion, Illumina miseq*), matching 1336 (0.6%) of the articles in LitCovid. We applied the filters to an updated version of LitCovid—–359,630 PubMed publications—–and randomly sampled 245 of the matching full-text articles for our final manual annotation (Figure 1).

**Figure 1:**
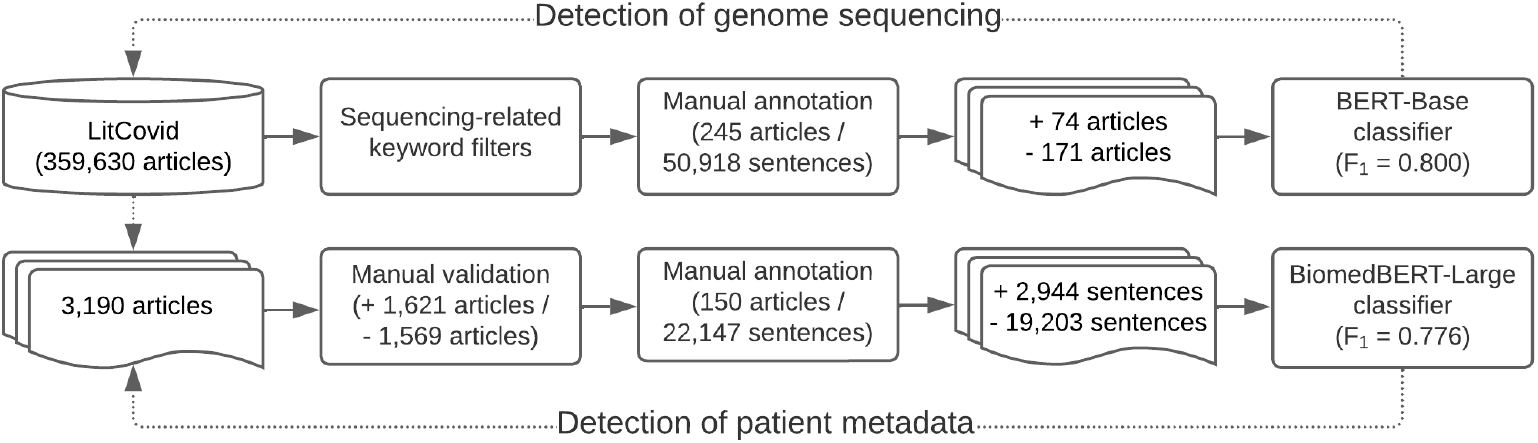
Workflow of natural language processing methods for detecting reports of SARS-CoV-2 sequencing and associated patient metadata in published articles.

### 2.2. Detection of genome sequencing

#### 2.2.1. Annotation

To prepare the 245 articles for manual annotation, we used the BioC [10] Python library to decode their BioC XML format, and then the spaCy Python library to segment the articles into 50,918 sentences. Sentence-level annotation allowed us to emphasize inclusion criteria that may not be the article’s primary focus and, thus, may be challenging to detect or explain by modeling larger units of text. We re-encoded the XML format of the sentences and imported them into the INCEpTION annotation platform [11]. Guidelines were developed to help annotators—four undergraduate students studying biomedical informatics (NK, ELW, HS, and PS)—determine whether the articles reported generating SARS-CoV-2 sequences—in particular, generating original sequences for the present study, as opposed to using sequences generated in a prior study—and identify specific sentences that provided evidence of such. Among the 245 articles, 75 were independently annotated by two of the annotators, with an inter-annotator agreement (F_1_-score) of 1.0 for identifying articles that reported generating SARS-CoV-2 sequences and 0.61 for identifying evidential sentences. Disagreements were independently resolved by one of the authors (AE). Among the 245 articles, 74 (30%) reported generating SARS-CoV-2 sequences and 171 (70%) did not (Figure 1). Among the 50,918 sentences, 357 (0.70%) provided evidence of generating SARS-CoV-2 sequences (“positive” class), while 50,561 (99.3%) did not (“negative” class)—a highly imbalanced dataset for supervised classification of sentences [12].

#### 2.2.2. Classification

We split the 245 annotated articles into three random sets: 147 articles (31,885 sentences) for training, 49 articles (9017 sentences) for validation, and 49 articles (10,016 sentences) as held-out test data. Although the classifiers were trained at the sentence level, we were primarily interested in evaluating their performance at the article level because our downstream task was to search for patient metadata within entire articles that reported generating SARS-CoV-2 sequences. Thus, we split the sets at the article level (i.e., there were no sentences from the same articles across the training, validation, and test sets) to be able to model this evaluation. We evaluated the classifiers based on the F_1_-score for the “positive” class: *F*_*1*_*-score = 2 * (precision * recall) / (precision + recall)*. While optimizing the F_1_-score, we were more interested in achieving a higher recall—*recall = true positives / (true positives + false negatives)*—than precision—*precision = true positives / (true positives + false positives)*—in order to retain a large number of true positive articles for downstream processing, especially given their relative sparsity. In our evaluation of the classifiers, we considered an article to be a true positive if it contained at least one true positive sentence.

As a baseline, we trained and evaluated a support vector machine (SVM) classifier, using the LibSVM [13] implementation in Weka. We pre-processed the sentences by normalizing URLs and numbers, removing non-alphanumeric characters and stopwords, and lowercasing and stemming [14] the text. We extracted n-grams (n=1-3) as features in a bag-of-words representation. We used the radial basis function (RBF) kernel and set the *cost* parameter at *c*=32 and, to address the class imbalance [12], the *weights* parameter at *w* =1 for the “negative” (i.e., majority) class and *w* =30 for the “positive” (i.e., minority) class, using the validation set to optimize these parameters. We also trained and evaluated a deep neural network classifier by fine-tuning the BERT-Base-Uncased [15] pre-trained model in the Hugging Face [16] library (Figure 1). We used 20 epochs, a batch size of 16, and a learning rate of 2e-5. We used the validation set to evaluate the models after each epoch, and re-loaded the best-performing model based on the validation set to evaluate its performance on the held-out test set. To address the class imbalance, we performed an additional experiment with the BERT-based classifier in which we under-sampled the “negative” (i.e., majority) class in the training set by removing sentences that were not similar to any of the sentences in the “positive” (i.e., the minority) class. We used similarity metrics to compare the sentences’ vector representations based on a pre-trained sentence transformer model [17].

While keyword filters were necessary initially to curate a set of articles for manual annotation, an evaluation of the classifier’s performance based on the keyword-matched test set would not necessarily reflect its performance in general. To evaluate the generalizability of the classifier, we deployed it on the sentences of all 117,327 full-text articles that were available in LitCovid [9] at this point in our study (Figure 1). We evaluated precision by manually annotating true and false positives among the sentences that the classifier predicted as providing evidence that the study had generated SARS-CoV-2 sequences. Given that 99.3% of the annotated sentences in the keyword-matched articles did not provide evidence of generating SARS-CoV-2 sequences, it would not have been feasible to evaluate recall by manually annotating even a representative sample of the sentences as true or false negatives. Instead, we identified a sample of 35 articles in LitCovid that were associated with GenBank [3] accession numbers but did not match any of our keywords, and we evaluated the articles that the classifier predicted as true positives and false negatives, in addition to the 12 articles in the keyword-matched test set that reported generating SARS-CoV-2 sequences.

### 2.3. Detection of patient metadata

#### 2.3.1. Annotation

We manually validated the articles in LitCovid [9] for which the deployed classifier predicted the presence of sentences that provided evidence of generating SARS-CoV-2 sequences, and randomly sampled 150 of them for further manual annotation using the INCEpTION platform [11] (Figure 1). Guidelines were developed to help annotators—two of the authors (AZK and KO) and an undergraduate student studying biomedical informatics (NK)—identify sentences that reported patient metadata associated with the SARS-CoV-2 sequences, including age, sex, race/ethnicity, symptoms, disease severity, viral load, duration of infection, lab results, vital signs, treatments, hospitalization, outcomes, comorbidities, risk factors, vaccination status, place of residence, geographic location of sample collection, and travel history. For studies in which sequences were generated for only a subset of the samples, we included patient metadata only if we could determine its association with samples that underwent sequencing. All 22,147 sentences in the 150 articles were independently annotated by two of the annotators, with an inter-annotator agreement (F_1_-score) of 0.75 for identifying sentences that reported patient metadata. Disagreements were resolved through discussion. Among the 22,147 sentences, 2944 (13.3%) reported patient metadata (“positive” class) and 19,203 (86.7%) did not (“negative” class)—again, an imbalanced dataset for supervised classification [12] (Figure 1).

#### 2.3.2. Classification

We randomly split the 22,147 annotated sentences into 15,504 (70%) sentences for training, 2214 (10%) sentences for validation, and 4429 (20%) sentences as held-out test data, stratified based on the distribution of the binary classes. While the training, validation, and test sets for the first classifier in our pipeline were split at the article level, we split the sets for this down-stream classifier at the sentence level in order to reduce a potential sampling bias; namely, some types of patient metadata were sparse and concentrated in a small subset of articles, so it would have been more likely for them not to be represented in the test set if it were split at the article level. We trained and evaluated deep neural network classifiers by fine-tuning BERT-based models in the Hugging Face [16] library that were pre-trained on corpora in general domains (DistilBERT-Base [18], BERT-Base [15], and RoBERTa-Large [19]), biomedical domains (BioBERT [20], BiomedBERT-Large-Abstract [21], and BiomedBERT-Base-Abstract-Fulltext [22]), and, more specifically, COVID-19 domains (COVID-SciBERT [23] and COVID-Twitter-BERT [24]) (Figure 1). We used 5 epochs, a batch size of 8, and a learning rate of 1e-5. We also fine-tuned the Llama-3-8B [25] large language model (LLM), using 10 epochs and a learning rate of 5e-5. To address the class imbalance [12], we performed additional experiments in which we randomly under-sampled the “negative” (i.e., majority) class and used the scikit-learn [26] Python library to automatically compute class weights, which assign more importance to, and a higher penalty for misclassifying, the “positive” class.

In addition to fine-tuning an LLM as a machine learning approach, we also experimented with prompting an LLM (Llama-3-70B [25]) as a generative artificial intelligence (AI) approach—that is, instructing the foundation model to detect sentences that reported patient metadata without further training. We developed and evaluated various prompts for an ablation study, with the *temperature* parameter set at 0.1 and the *top p* parameter at 0.9. As a zero-shot baseline (i.e., without providing any examples for the LLM), our first prompt simply defined patient metadata and then instructed the LLM to answer whether or not each input sentence reported or referred to (e.g., in tables, figures, or supplementary material) patient metadata, explicitly restricting the response to *yes* or *no*. Second, we used role-based prompting, adding the following “system role” to the zero-shot prompt in order to provide additional context about the perspective from which the LLM should respond: *You are a genomic epidemiologist who is reviewing scientific literature about SARS-CoV-2 genome sequencing*. Third, we used few-shot prompting, or in-context learning, adding a small number of diverse sentences from the training set as examples that reported or referred to patient metadata. Fourth, we combined few-shot prompting with chain-of-thought prompting [27], providing reasoning for each example, such as: *This sentence refers to a table in supplementary material that includes patient metadata about age, comorbidities, and race/ethnicity*. In our final prompt, we elicited reasoning by the LLM prior to answering the question, and instructed the LLM to provide that reasoning to support its *yes* or *no* response.

## 3. Results

### 3.1. Detection of genome sequencing

#### 3.1.1. Test set evaluation

Based on the 49 keyword-matched articles (10,016 sentences) in the held-out test set, the SVM [13] classifier achieved an F_1_-score of 0.370 (precision = 0.329 and recall = 0.422) and the BERT-based [15] classifier achieved an F_1_-score of 0.480 (precision = 0.492 and recall = 0.469) for detecting sentences that provided evidence of generating SARS-CoV-2 sequences. Experimenting with an under-sampled training set to address the class imbalance [12], the BERT-based classifier achieved an F_1_-score of 0.453 (precision = 0.453 and recall = 0.453). Despite the classifiers’ moderate performance at the sentence level, the performance of the BERT-based model that was fine-tuned using the full training set achieved an F_1_-score of 0.800 (precision = 0.667 and recall = 1.0) at the article level. Recall increased from the sentence level to the article level because the classifier detected at least one true positive sentence in all 12 articles that reported generating SARS-CoV-2 sequences, even though there were also false negative sentences in these 12 articles. Precision increased from the sentence level to the article level because there were more false positive sentences among these 12 true positive articles than among the 37 true negative articles.

#### 3.1.2. LitCovid evaluation

Among the 117,327 full-text articles that were available in LitCovid [9] at the time of deployment, the BERT-based classifier predicted that 3190 of them contained sentences that provided evidence of generating SARS-CoV-2 sequences. We manually validated these 3190 articles, determining that 1621 (50.8%) were true positives and 1569 (49.2%) were false positives. Among these 1621 true positive articles, 492 (30.4%) did not match the keywords used to filter the annotated dataset and, thus, would not have been retrieved using a traditional keyword-based literature search. As expected, the classifier’s article-level precision decreased—from 0.667 to 0.508—when it was deployed on sentences in articles that did not match the keywords used to select the articles in the training set. Among 47 articles in LitCovid that reported generating SARS-CoV-2 sequences—12 in the test set and 35 that were associated with GenBank [3] accession numbers but did not match any of the keywords—–the BERT-based classifier predicted true positive sentences in 40 (85.1%) of them. As expected, the classifier’s article-level recall also decreased—–from 1.0 to 0.851—when it was deployed on sentences in articles that were not represented in the training set. Nonetheless, recall remained high, suggesting that the 1621 articles that we manually validated represented most of the full-text articles in PubMed that reported generating SARS-CoV-2 sequences at the time of this study. Given the article-level recall of the BERT-based classifier, experimenting with additional pre-trained models would have been superfluous.

#### 3.1.3. Error analysis

We conducted an error analysis of one false positive sentence per article (Table 1). Among the 1,569 false positive sentences, 645 (41.1%) were unrelated to sequencing but included words that were frequently used to report sequencing (e.g., *obtained, assembled*); 273 (17.4%) reported using prior sequences rather than generating original sequences; 123 (7.8%) reported retrieving sequences, rather than generating sequences; 90 (5.7%) reported a study sample; 84 (5.4%) included technical terms related to sequencing (e.g., *consensus*) that were not being used in a sequencing context; 69 (4.4%) reported generating sequences that were either not SARS-CoV-2, whole genome, or human; 51 (3.3%) reported a literature search; 8 (0.5%) reported an *in silico* study; 6 (0.4%) reported negative results of sequencing; and 220 (14.0%) were other errors. Many of the false positives—particularly those that reported prior sequencing or retrieving sequences—required the annotators to rely on surrounding sentences to distinguish them.

**Table 1:** False positives (N=1569) of a BERT-based classifier for detecting sentences that reported generating SARS-CoV-2 sequences among 117,327 full-text PubMed articles in the LitCovid collection.

| Error type | n, % | Example |
| --- | --- | --- |
| Sequence report word | 645 (41.1%) | We obtained COVID-19 country-wide data from the Johns Hopkins Center for Systems Science and Engineering. |
| Prior sequencing | 273 (17.4%) | A new, human coronavirus (HCoV) was isolated from these cases and identified as a betacoronavirus and provisionally named 2019 novel corona virus (2019-nCoV) using next-generation sequencing technology. |
| Retrieving sequences | 123 (7.8%) | Whole-genome sequence (WGS) or specific gene sequence data of SARS-CoV-2 were collected from NCBI Nucleotide GenBank and http://www.GISAID.org. |
| Study sample | 90 (5.7%) | In this study, we collected 334 confirmed COVID-19 patients including 212 still in hospital with nucleic acid test positive on halfway for SARS-CoV-2 and 122 discharged from hospital from Feb 27, 2020, to Mar 1, 2020, in Wuhan Union hospital, a designated hospital for treating COVID-19. |
| Irrelevant context | 84 (5.4%) | Then, we applied consensus clustering to the results of ensemble classification. |
| Irrelevant sequencing | 69 (4.4%) | The three dog samples were sent to Macrogen (South Korea) for whole genome sequencing. |
| Literature search | 51 (3.3%) | To find relevant primitive articles, we performed a comprehensive search in databases, involving Medline via PubMed, EMBASE and Web of Science applying the following terms: ‘COVID-19’, ‘SARS-CoV-2’, ‘Wuhan virus’, [...]. |
| In silico study | 8 (0.5%) | We simulated SARS-CoV-2 genomes with SANTA-SIM, using the consensus sequence WuhanHu-1 as input sequence available at https://www.ncbi.nlm.nih.gov/nuccore/MN908947.3. |
| Negative results | 6 (0.4%) | All re-positive case samples were unsuccessfully sequenced. |
| Other | 220 (14.0%) | – |

### 3.2. Detection of patient metadata

#### 3.2.1. Evaluation of fine-tuned models

Based on the 4429 sentences in the held-out test set, fine-tuned models that were pre-trained on corpora in biomedical domains (BioBERT [20], BiomedBERT-Large-Abstract [21], and BiomedBERT-Base-Abstract-Fulltext [22]) or, more specifically, COVID-19 domains (COVID-SciBERT [23] and COVID-Twitter-BERT [24]) outperformed those that were pre-trained on corpora in general domains (DistilBERT-Base [18], BERT-Base [15], and RoBERTa-Large [19]) for detecting sentences that reported patient metadata associated with SARS-CoV-2 sequences, including the Llama-3-8B [25] LLM (Table 2). The classifier based on BiomedBERT-Large-Abstract—a model pre-trained on PubMed abstracts—achieved the highest F_1_-score (0.776). It marginally outperformed classifiers based on BiomedBERT-Base-Abstract-Fulltext (0.767)—a smaller model pre-trained on PubMed abstracts and PubMed Central full-text articles—and COVID-Twitter-BERT (0.766)—a model pre-trained on Twitter posts related to COVID-19. We performed additional experiments for these latter three classifiers to address the class imbalance [12]; however, while under-sampling combined with class weights improved recall for all three classifiers, the F_1_-score did not improve (Table 3).

**Table 2:** Precision (P), recall (R), and F_1_-score (F_1_) of fine-tuned classification models for detecting sentences in PubMed articles that reported patient metadata associated with SARS-CoV-2 sequences.

| **Model** | **P** | **R** | **$F_1$** |
| --- | --- | --- | --- |
| DistilBERT-Base | 0.760 | 0.698 | 0.727 |
| BERT-Base | 0.733 | 0.718 | 0.726 |
| RoBERTa-Large | 0.730 | 0.650 | 0.688 |
| BioBERT | 0.740 | 0.744 | 0.742 |
| BiomedBERT-Large-Abstract | 0.743 | 0.812 | 0.776 |
| BiomedBERT-Base-Abstract-Fulltext | 0.740 | 0.796 | 0.767 |
| COVID-SciBERT | 0.775 | 0.677 | 0.723 |
| COVID-Twitter-BERT | 0.752 | 0.779 | 0.766 |
| Llama-3-8B | 0.691 | 0.660 | 0.675 |

**Table 3:** Precision (P), recall (R), and F_1_-score (F_1_) of fine-tuned classification models for detecting sentences in PubMed articles that reported patient metadata associated with SARS-CoV-2 sequences, with approaches for addressing class imbalance.

| **Model** | **P** | **R** | **F<sub>1</sub>** |
| --- | --- | --- | --- |
| BiomedBERT-Large-Abstract |  |  |  |
| Baseline | 0.743 | 0.812 | 0.776 |
| Under-sampling | 0.749 | 0.749 | 0.749 |
| Class weights | 0.769 | 0.767 | 0.768 |
| Under-sampling + class weights | 0.694 | 0.815 | 0.749 |
| BiomedBERT-Base-Abstract-Fulltext |  |  |  |
| Baseline | 0.740 | 0.796 | 0.767 |
| Under-sampling | 0.708 | 0.784 | 0.744 |
| Class weights | 0.768 | 0.764 | 0.766 |
| Under-sampling + class weights | 0.678 | 0.820 | 0.743 |
| COVID-Twitter-BERT |  |  |  |
| Baseline | 0.752 | 0.779 | 0.766 |
| Under-sampling | 0.707 | 0.727 | 0.717 |
| Class weights | 0.761 | 0.740 | 0.750 |
| Under-sampling + class weights | 0.717 | 0.803 | 0.757 |

#### 3.2.2. Evaluation of LLM prompting

Based on the 4429 sentences in the held-out test set, zero-shot prompting of an LLM (Llama-3-70B [25]) achieved an F_1_-score of 0.482 (Table 4). Adding a “system role” to the zero-shot prompt improved the F_1_-score to 0.494. While few-shot prompting alone did not substantially improve the F_1_-score of role-based prompting (0.497), few-shot prompting combined with chain-of-thought prompting improved the F_1_-score to 0.558. Instructing the LLM to provide reasoning to support its *yes* or *no* response did not improve the F_1_-score (0.551), achieving the highest overall precision among the prompts (0.532) but substantially reducing recall (0.491). Our best-performing generative AI approach, however, was outperformed by all of our machine learning-based classifiers that used manually annotated training data to fine-tune pre-trained models. While prompting an LLM would outperform the fine-tuned BiomedBERT-Large-Abstract [21] model if nearly no training data were available, the fine-tuned model only required approximately 5% of our training set to begin outperforming the LLM (Figure 2).

**Table 4:** Precision (P), recall (R), and F_1_-score (F_1_) of LLM (Llama-3-70B) prompting for detecting sentences in PubMed articles that reported patient metadata associated with SARS-CoV-2 sequences.

| Prompt | P | R | F<sub>1</sub> |
| --- | --- | --- | --- |
| Zero-shot | 0.448 | 0.521 | 0.482 |
| Zero-shot + role | 0.416 | 0.608 | 0.494 |
| Role + few-shot | 0.508 | 0.486 | 0.497 |
| Role + few-shot + chain-of-thought | 0.510 | 0.616 | 0.558 |
| Role + few-shot + chain-of-thought + reasoning | 0.532 | 0.491 | 0.511 |

**Figure 2:**
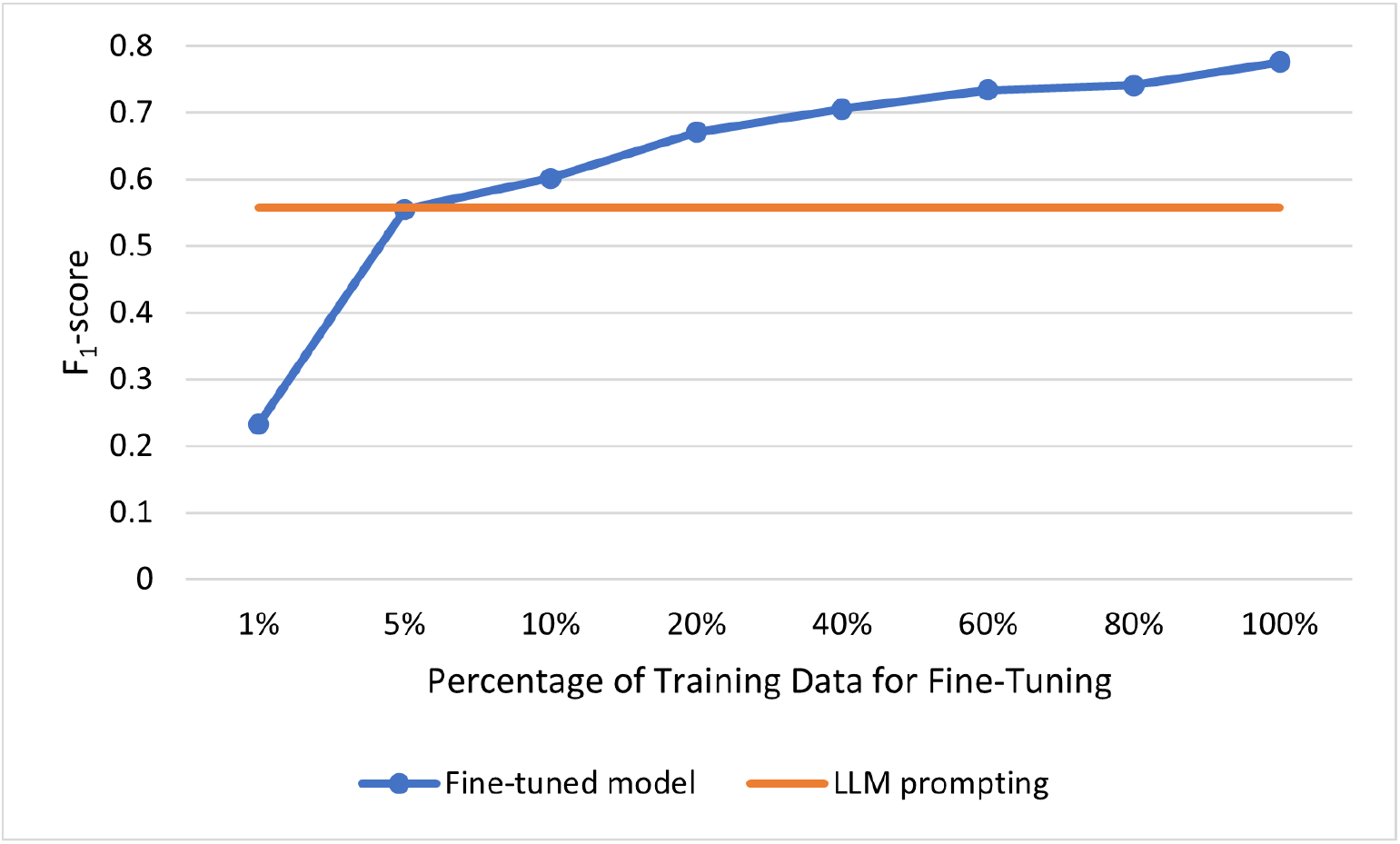
F_1_-scores of a fine-tuned model (BiomedBERT-Large-Abstract), using varying amounts of manually annotated training data, and LLM (Llama-3-70B) prompting for detecting sentences in PubMed articles that reported patient metadata associated with SARS-CoV-2 sequences.

#### 3.2.3. Error analysis

We conducted an error analysis focusing on a comparison of the test set predictions between the fine-tuned BiomedBERT-Large-Abstract [21] model and the Llama-3-70B [25] LLM. Although instructing the LLM to provide reasoning resulted in a lower F_1_-score, we used this prompt for error analysis because the elaborated responses provided valuable insight for understanding its misclassifications and, in turn, potentially improving the prompts.

Among the 225 sentences that were predicted as false negatives by the LLM but correctly classified by the fine-tuned model, 183 (81.3%) reported a geographic location associated with SARS-CoV-2 sequences, 186 (82.7%) of the LLM’s responses emphasized that the sentence was not about patients (e.g., *patient-specific, individual patients, patients themselves*), and 189 (84%) of the LLM’s responses suggest that the LLM may have invoked a preconceived definition of patient metadata. For example, the following sentence reported a geographic location—the Hisar district in Haryana, India—in which samples that underwent sequencing were collected: *On the onset of the second wave of COVID-19 (April/May 2021) in India, we again attempted virus isolation from the samples received from the Hisar district of Haryana*. Although the LLM affirmed that the sentence did provide information about the geographic location of sample collection, it disregarded that such information had been explicitly defined as patient metadata and illustrated with examples in the prompt, focusing instead on types of metadata that more directly characterize patients: *No. This sentence does not report or refer to patient metadata. It provides information about the timing and location of sample collection, but it does not mention any specific patient characteristics, such as age, sex, symptoms, disease severity, or other metadata. The sentence is describing the context of the sample collection, rather than providing information about individual patients*. Among the 36 sentences that were predicted as false negatives by the fine-tuned model but correctly classified by the LLM, 30 (83.3%) explicitly referred to patients (e.g., *patients, individuals, residents, cases, persons, subjects*), and 25 (69.4%) included patient metadata other than geographic location.

While the fine-tuned model performed better than the LLM at detecting sentences that reported geographic location, over-generalization was the primary source of the fine-tuned model’s false positives. Among the 104 sentences that were predicted as false positives by the fine-tuned model but correctly classified by the LLM, 76 (73.1%) included a geographic location that was not reported within the context of SARS-CoV-2 sequences. As with the false negatives, among the 193 sentences that were predicted as false positives by the LLM but correctly classified by the fine-tuned model, 50 (25.9%) of the LLM’s responses suggest that the LLM disregarded the definition and examples of patient metadata in the prompt. For the false negatives, the LLM excluded particular types of patient metadata, whereas, for the false positives, the LLM either misconstrued particular types of patient metadata or included particular types of patient metadata that were not provided in the prompt. For example, the following sentence did not report patient metadata as defined and illustrated in the prompt: *The majority (15/18) of patients with a positive conjunctival sample were healthcare workers*. The LLM, however, added “occupation” to the definition of patient metadata and misconstrued a positive SARS-CoV-2 sample as patient metadata about a lab result, despite the example in the prompt: *Yes. This sentence reports patient metadata about occupation (healthcare workers) and a lab result (positive conjunctival sample)*. While 161 (83.4%) of the 193 false positives did include types of patient metadata that were defined in the prompt, the patient metadata reported in 116 (60.1%) of them were not clearly associated with SARS-CoV-2 samples that underwent sequencing, and 51 (26.4%) of the sentences did not report specific values, such as merely reporting that patient metadata were collected: *Associated epidemiological data, such as symptoms, travel history and municipality of residency, were collected from medical records accompanying the collected samples provided by IOM/FUNED*.

## 4. Discussion

While the utility of genomic epidemiology for SARS-CoV-2—that is, the use of SARS-CoV-2 sequences to understand transmission patterns—depends on metadata such as the patients’ age, sex, geographic location, travel history, symptoms, and clinical outcomes, much of this metadata had been digitally disconnected from sequence databases and “locked” in the unstructured text among more than 350,000 published articles relevant to COVID-19 at the time of this study. The results of this study demonstrate that our NLP methods can be used to facilitate the large-scale detection of patient metadata associated with reports of SARS-CoV-2 sequencing in these published articles. Although this study focused on mining articles relevant to COVID-19 in particular, our NLP pipeline is potentially generalizable across viruses, since our methodology—the collection, annotation, and classification of all sentences in samples of full-text articles—was not limited to select genome sequences or types of patient metadata, but rather was based on modeling the broad detection of such reports in general.

More generally, the results of this study suggest that fine-tuning BERT-based models using manually annotated training data may outperform LLM prompting for classification tasks involving complex concepts, such as *patient metadata*. Nonetheless, our error analysis provided valuable insight for potentially improving the performance of the prompts in future work. For example, the prompts first defined patient metadata and then instructed the LLM to answer whether or not each input sentence reported or referred to patient metadata, requiring the LLM to understand that the instruction referred to the provided definition. Because the LLM’s responses suggest that the definition was disregarded and that LLM focused its attention on “patient metadata” in the instruction, we will experiment with removing this terminology and incorporating the types of patient metadata directly into the instruction, rather than predefining patient metadata as a referent. In doing so, we will also clarify particular types of patient metadata that the LLM had misconstrued (e.g., lab results). Given that few-shot prompting combined with chain-of-thought prompting improved performance using examples of sentences that reported or referred to patient metadata, we will also experiment with adding examples of sentences that did not report or refer to patient metadata, and providing reasoning such as: *This sentence reports patient metadata associated with sequences that were retrieved from a database, not sequences that were generated for this study*.

The primary limitation of this study is that our evaluation of the classifiers for detecting reports of patient metadata was based only on the manually validated output of the first classifier in our pipeline—that is, true positive articles that reported generating SARS-CoV-2 sequences. By manually excluding the false positive articles, our evaluation of the downstream classifiers did not represent their performance for a fully automated pipeline. Because articles that reported generating SARS-CoV-2 sequences were relatively sparse, we decided to prioritize high recall—a trade-off that increased the number of false positive articles. Given that these false positive articles would likely include irrelevant reports of patient metadata and, thus, decrease the performance of the downstream classifiers, we decided to take a human-in-the-loop approach. Despite this limitation, our approach would nonetheless enable the efficient use of published articles for genomic epidemiology, reducing the number of full-text articles in this study (117,327) by more than 97% (3190) for manual validation. Furthermore, the articles that we manually validated were a subset of an extensive collection of articles that had been published over the course of several years and then retrospectively downloaded for this study. In contrast, the input to our human-in-the-loop approach would likely be much smaller for particular applications of genomic epidemiology in real-time.

## 5. Conclusion

In this paper, we presented NLP methods to facilitate the large-scale detection of patient metadata associated with reports of genome sequencing in published articles, advancing the utility and efficiency of genomic epidemiology for timely public health responses to virus outbreaks. In future work, we will further assess the generalizability of our methodology across viruses and use our error analysis to inform experiments for improving the performance of LLM prompting for complex concepts.

## Data Availability

The annotation guidelines, data, and LLM prompts produced in the present study are available upon reasonable request to the authors.

## Acknowledgments

This work was supported by the National Institute of Allergy and Infectious Diseases (R01AI164481). The content is solely the responsibility of the authors and does not necessarily represent the official views of the National Institutes of Health. The authors thank Nicole Kaiser, Ethan Leiter-Weintraub, Haris Siddiq, and Peter Skidmore for contributing to annotation.

